# Lack of trust and social media echo chambers predict COVID-19 vaccine hesitancy

**DOI:** 10.1101/2021.01.26.21250246

**Authors:** Will Jennings, Gerry Stoker, Hannah Willis, Viktor Valgardsson, Jen Gaskell, Daniel Devine, Lawrence McKay, Melinda C. Mills

**Author notes:** Corresponding authors: Will Jennings and Melinda C. Mills.

## Abstract

As COVID-19 vaccines are rolled out across the world, there are growing concerns about the role that trust, belief in conspiracy theories and spread of misinformation through social media impact vaccine hesitancy. We use a nationally representative survey of 1,476 adults in the UK between December 12 to 18, 2020 and five focus groups conducted in the same period. Trust is a core predictor, with distrust in vaccines in general and mistrust in government raising vaccine hesitancy. Trust in health institutions and experts and perceived personal threat are vital, with focus groups revealing that COVID-19 vaccine hesitancy is driven by a misunderstanding of herd immunity as providing protection, fear of rapid vaccine development and side effects, belief the virus is man-made and related to population control. Particularly those who obtain information from relatively unregulated social media sources such as YouTube that have recommendations tailored by watch history are less likely to be willing to become vaccinated. Those who hold general conspiratorial beliefs are less willing to be vaccinated. Since an increasing number of individuals use social media for gathering health information, interventions require action from governments, health officials and social media companies. More attention needs to help people understand their own risks, unpack complex concepts and fill knowledge voids.

## Introduction

Governments are rapidly mobilising vaccines against COVID-19 (1), with success relying on sufficient uptake. Yet there is a rise in vaccine hesitancy linked to loss of trust, complacency and misinformation (2, 3). Trust is crucial to ensure compliance to public health measures (4). But governments, experts and the media have needed to communicate uncertain and even reversals in advice, eroding public trust (5). COVID-19 is not only a pandemic, but an ‘infodemic’ of complex and dynamic information – both factual and incorrect. This can generate vaccine hesitancy, which the WHO listed as one of the top 10 threats to global health in 2019. But who does the public trust and does this depend on where they acquire their information? The growth in internet use and reliance on social media sources such as YouTube, Facebook, Twitter and TikTok has changed the landscape of information gathering, with 72% of Americans and 83% of Europeans using the internet as a source for health information (6). Conspiracy and anti-vaxx beliefs and low trust in institutions is associated with a greater reliance on social media for health information, but until now primarily only with small selective samples (e.g., MTurk) (7, 8). To empirically inform these urgent issues, we present the results of a survey fielded during the first vaccine roll-out in the UK between December 12 to 18, 2020 on a nationally representative sample of 1,476 adults, complemented with five focus groups conducted in roughly the same period (see SI).

### Trust, threat, and information sources

We test three hypotheses. First, we contend that multiple facets of trust are crucial in understanding vaccine uptake (4, 5, 9). We hypothesise that trust in government and a positive view of the government’s handling of the crisis will predict higher vaccine willingness, while vaccine distrust and mistrust and distrust of government predicts greater hesitancy (see SI for measures, full analyses).

Trust is confidence in the action of others, mistrust measures vigilance in whether actors or information are trustworthy and distrust denotes a negative orientation towards institutions or actors (4). A recent survey in England found those endorsing conspiracy theories were less likely to adhere to government guidelines and had a general distrust in institutions (10). Individuals may not trust the government, but more willing to ‘follow the science’ and trust scientific or health experts. We therefore predict those with higher levels of trust in health institutions and experts will exhibit higher vaccine willingness (8).

Second, we hypothesise that those with higher collective social trust and perceived personal threat from COVID-19 are less vaccine hesitant. Social trust enables the collective action needed to achieve sufficient population vaccination levels, with social capital positively associated with health (11).

Since deaths from COVID-19 are concentrated in higher ages and risk groups (12), public discourse has been centred around ‘vulnerable’ groups and herd immunity (13). If risks are perceived as low, it translates into lower vaccination intentions (14, 15).

Third, we expect that consumers of social media are more likely to be vaccine hesitant than those of traditional media sources (TV, newspaper, radio). Holding general conspiracy or COVID-19 misinformation beliefs will likewise lower vaccine willingness. The main sources of vaccine misinformation are on social media. An analysis of 1,300 Facebook pages during the 2019 measles outbreak found anti-vaxx pages grew by 500%, compared to 50% of pro-vaccine pages (16).

Individuals can also end in echo chambers; once a YouTube user develops a watch history, a filter bubble tailors their Top 5 and Up-Next recommendations, with watching videos promoting vaccine misinformation leading to more misinformed recommendations (17).

We also anticipate socio-political-demographic variation by digital disparities in information seeking by younger, more educated and higher socio-economic status individuals (6, 18). Political conservatives are more likely to believe in vaccine conspiracies (8). An analysis of popular anti-vaxx Facebook pages found the majority (72%) were mothers (19).

## Results

We analyse vaccine willingness by asking “If a vaccine for COVID-19 were available to me, I would get it”, dichotomised into strongly agree or tend to agree (71%) versus those who strongly or tend to disagree, neither or are unsure (29%). 49% strongly agreed they would get the vaccine, 22% tended to agree they would get it, 11% neither agreed or disagreed, 7% tended to disagree and 7% strongly disagreed (with 5% indicating don’t know).

Our results are in Figure 1 and Table 1 with detailed focus group results in the SI. We find evidence for H1, H2 and nuanced findings for H3. For H1, those expressing the highest levels of vaccine distrust have a 10% probability of vaccine willingness. The effect size is not surprising given the proximity to our dependent variable. Those who mistrust government are more hesitant, with the highest level of mistrust having a 25% probability of vaccine willingness. Those with the highest level of trust in health institutions are twice as likely to express vaccine willingness compared to the lowest level of trust. We also find a significant positive association for trust in experts.

**Tab 1.** Logistic regression estimates of vaccine willingness, odds ratios

|  | (1) | (2) | (3) | (3*) |
| --- | --- | --- | --- | --- |
| *Trust* |  |  |  |  |
| Social trust | 1.273<br>(0.923 - 1.754) | 1.254<br>(0.901 - 1.745) | 1.228<br>(0.880 - 1.714) | 1.205<br>(0.852 - 1.705) |
| Trust: government | 0.696<br>(0.242 - 2.005) | 0.705<br>(0.240 - 2.075) | 0.696<br>(0.234 - 2.070) | 0.751<br>(0.243 - 2.324) |
| Mistrust: government | 0.400<br>(0.164 - 0.977)* | 0.381<br>(0.153 - 0.949)* | 0.395<br>(0.157 - 0.996)* | 0.349<br>(0.133 - 0.913)* |
| Distrust: government | 2.420<br>(0.956 - 6.125) | 2.363<br>(0.896 - 6.227) | 2.180<br>(0.817 - 5.818) | 2.665<br>(0.958 - 7.415) |
| Trust: health organisations | 6.154<br>(2.735 - 13.846)*** | 6.294<br>(2.715 - 14.592)*** | 6.019<br>(2.569 - 14.105)*** | 6.218<br>(2.560 - 15.104)*** |
| Trust: media | 1.394<br>(0.972 - 2.001) | 1.306<br>(0.901 - 1.894) | 1.349<br>(0.926 - 1.965) | 1.428<br>(0.965 - 2.112) |
| Trust: experts | 1.958<br>(0.810 - 4.736) | 1.716<br>(0.697 - 4.225) | 1.718<br>(0.692 - 4.264) | 2.695<br>(1.013 - 7.171)* |
| *COVID-19/Vaccines* |  |  |  |  |
| Government handling of COVID-19 | 2.323<br>(1.020 - 5.292)* | 2.131<br>(0.920 - 4.939) | 2.204<br>(0.945 - 5.142) | 2.249<br>(0.939 - 5.389) |
| Perceived personal threat of COVID-19 | 2.329<br>(1.073 - 5.053)* | 2.221<br>(1.004 - 4.915)* | 2.216<br>(0.993 - 4.942) | 2.344<br>(1.016 - 5.405)* |
| Lockdown scepticism | 1.181<br>(0.446 - 3.128) | 0.906<br>(0.331 - 2.481) | 0.888<br>(0.322 - 2.447) | 1.026<br>(0.358 - 2.943) |
| Conspiracy beliefs | 0.307<br>(0.106 - 0.891)* | 0.292<br>(0.097 - 0.881)* | 0.294<br>(0.096 - 0.897)* | 0.279<br>(0.087 - 0.894)* |
| Vaccine distrust | 0.083<br>(0.043 - 0.159)*** | 0.090<br>(0.046 - 0.177)*** | 0.091<br>(0.046 - 0.180)*** | 0.088<br>(0.044 - 0.179)*** |
| COVID-19 misinformed | 0.445<br>(0.170 - 1.167) | 0.476<br>(0.178 - 1.275) | 0.490<br>(0.181 - 1.324) | 0.414<br>(0.143 - 1.199) |
| *Demographics* |  |  |  |  |
| Had COVID-19 | 1.257<br>(0.688 - 2.298) | 1.317<br>(0.712 - 2.434) | 1.280<br>(0.692 - 2.369) | 1.193<br>(0.641 - 2.222) |
| Female | 0.731<br>(0.528 - 1.012) | 0.797<br>(0.570 - 1.116) | 0.787<br>(0.554 - 1.116) | 0.797<br>(0.555 - 1.145) |
| Age | 14.897<br>(6.862 - 32.341)*** | 13.781<br>(6.126 - 31.002)*** | 10.341<br>(4.152 - 25.756)*** | 12.684<br>(4.848 - 33.189)*** |
| Graduate | 1.701<br>(1.239 - 2.334)** | 1.572<br>(1.133 - 2.181)** | 1.574<br>(1.127 - 2.198)** | 1.560<br>(1.098 - 2.215)* |
| Supports Conservative Party | 1.210<br>(0.806 - 1.818) | 1.202<br>(0.794 - 1.821) | 1.167<br>(0.768 - 1.773) | 1.146<br>(0.744 - 1.765) |
| *Media/information* |  |  |  |  |
| Information sources: online |  | 1.716<br>(1.028 - 2.864)* | 1.664<br>(0.985 - 2.809) | 1.860<br>(1.071 - 3.232)* |
| Information sources: people |  | 1.022<br>(0.535 - 1.951) | 1.028<br>(0.536 - 1.973) | 0.976<br>(0.492 - 1.938) |
| Information sources: traditional |  | 1.565<br>(0.666 - 3.676) | 1.557<br>(0.654 - 3.702) | 1.276<br>(0.514 - 3.167) |
| Social media use: Facebook |  |  | 1.131<br>(0.787 - 1.625) | 1.058<br>(0.726 - 1.541) |
| Social media use: Twitter |  |  | 1.331<br>(0.902 - 1.964) | 1.206<br>(0.799 - 1.819) |
| Social media use: Instagram |  |  | 0.832<br>(0.561 - 1.236) | 0.840<br>(0.557 - 1.268) |
| Social media use: Reddit |  |  | 1.004<br>(0.540 - 1.867) | 0.935<br>(0.493 - 1.774) |
| Social media use: Youtube |  |  | 0.672<br>(0.472 - 0.956)* | 0.669<br>(0.463 - 0.968)* |
| Social media use: Snapchat |  |  | 0.998<br>(0.567 - 1.755) | 1.006<br>(0.563 - 1.796) |
| Social media use: TikTok |  |  | 0.951<br>(0.512 - 1.764) | 0.915<br>(0.487 - 1.719) |
| Fact-checked an article online |  |  |  | 0.992<br>(0.481 - 2.043) |
| Posted political content online |  |  |  | 1.790<br>(0.752 - 4.260) |
| N | 1,348 | 1,316 | 1,316 | 1,261 |
| Pseudo R-squared | 0.28 | 0.29 | 0.29 | 0.30 |

Notes: \* $p < 0.05$ ; \*\* $p < 0.01$ ; \*\*\* $p < 0.001$ (95 per cent confidence intervals in parentheses)

**Fig 1.**
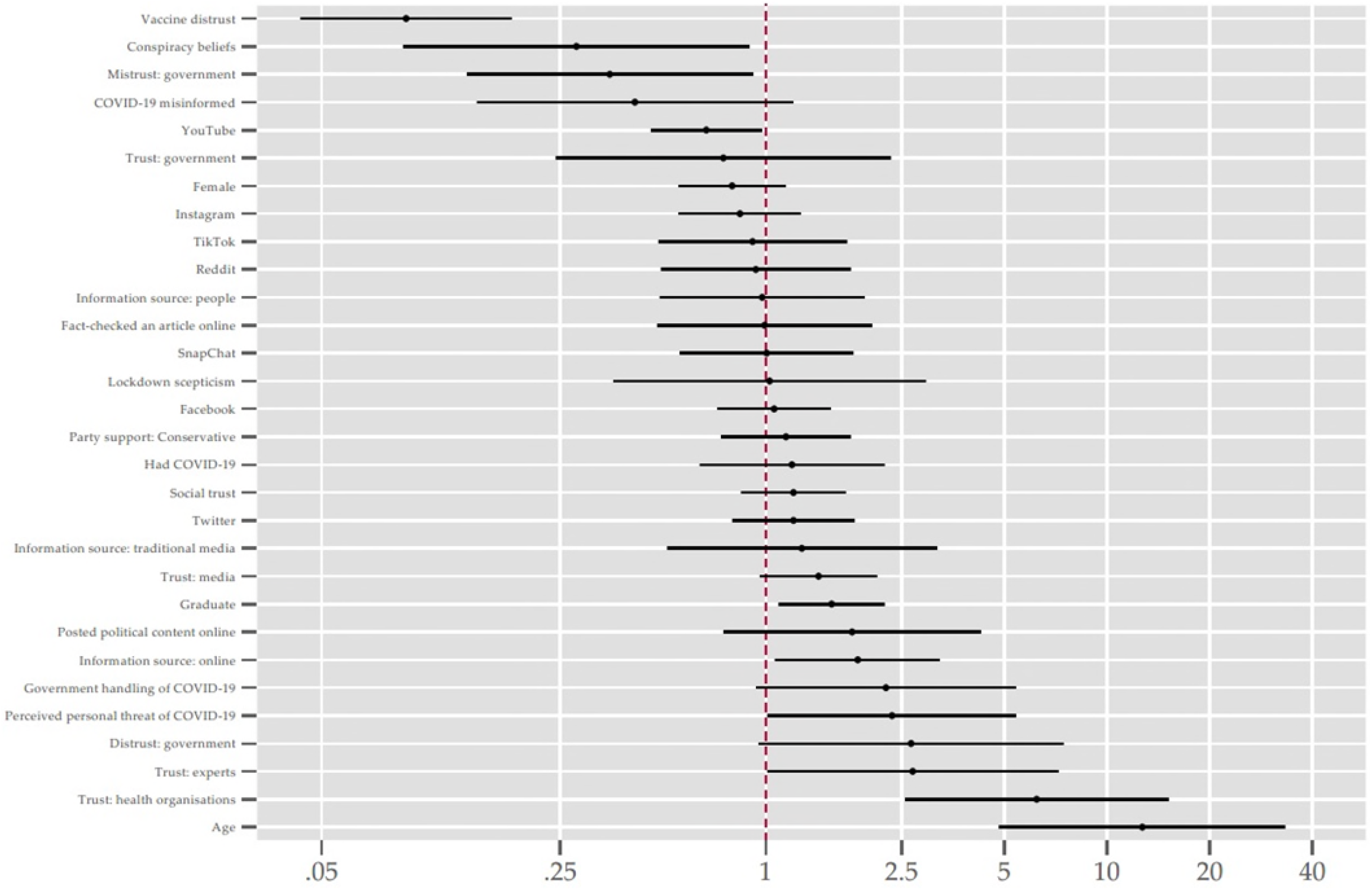
Odds ratios of determinants of vaccine willingness.

We do not find a significant effect for social trust in H2, but stratification across groups could result in divergent vaccine behaviour (see SI). A strong theme in the focus groups was scepticism over death rates, inconsistent COVID-19 policies in the ‘tier system’ and unfair burden and punishment of those in the North, which have higher levels of socio-economic deprivation.

Those who perceived COVID-19 as a personal threat were almost two and a half times more likely to express vaccine willingness than those who did not consider it a threat. A strong theme in the focus groups was that only the most vulnerable should get vaccinated, linked to ‘herd immunity’, which the government used in early messaging and was widely discussed pitting lockdowns versus natural herd immunity (13). This led them to believe that widespread infection would result in population immunity and little need for vaccination. Herd immunity is complicated and differs from the 70-80% vaccine herd immunity threshold, which is the proportion of the population required to block transmission, related to vaccine efficacy and immunity duration (20).

For H3, holding conspiracy beliefs is a significant predictor of vaccine hesitancy. Further, we find individuals who obtain more information from the internet are more willing to be vaccinated, but seeking online health information is widespread and heterogeneous. Only YouTube users were significantly less willing to be vaccinated, with a 45% probability of vaccine willingness. Instagram, TikTok and Snapchat users were more hesitant, but our sample size is too limited to draw conclusions. Facebook and particularly Twitter users have slightly higher odds of vaccine willingness, but not significant at the 95% confidence level. Our finding linking YouTube users to COVID-19 vaccine hesitancy is novel, but in line with research. A study of YouTube vaccine content found 65.5% of videos discouraged vaccine use focussing on autism, undisclosed risks, adverse reactions and mercury in vaccines (21). A 2017 analysis of 560 YouTube vaccine videos in Italy, found the majority were negative, linking vaccines with autism and serious side effects (22). Those who refused vaccines in the focus groups had low levels of trust in the government, believed the virus was man-made or a type of population control for certain groups. Individuals who are younger and with lower levels of education were also vaccine hesitant in our analysis.

## Discussion

We provide new evidence on how trust and information is linked with COVID-19 vaccine hesitancy, informing policy in key ways. Misinformation thrives where there is lack of trust in government, politics and elites. A broader lesson is the need for authorities to communicate truthfully and consistently. Over-promising, confusing messages and blame rather than solving problems are faults of government and politicians that are best minimised – especially during times of crisis.

Personal perceived threat remains pivotal. With increased vaccinations, a drop in infections and deaths, individuals perceive lower threat. Our focus group revealed complacency emerges from a misunderstanding of ‘herd immunity’. What may seem as irrational, conspiratorial judgements are often attempts to make sense of knowledge fragments accumulated during a fraught, complex and rapidly evolving crisis. The public use a ‘fast’ and frugal model of intuitive thinking, using a mix of short cuts and heuristics (23), which all should be taken in account in communications.

Since the internet and social media are key sources for health information, governments should establish an engaging web presence to fill knowledge gaps (3). Sites remain unregulated and not operating as ‘publishers’ forced to present balanced information, with misinformation or conspiracy theories quickly becoming ‘viral’. Advertisers can boycott their advertisements alongside harmful content (24), companies can check information, alter keyword searches and redirect to correct sources (3), ban overt conspiracy groups such as QAnon (3), balance viewpoints, flag misinformation or rapidly remove content. YouTube and Facebook removed ‘Plandemic’, but only after it was watched by millions (25). The most common sources of YouTube vaccine information were by non-expert individuals (21), suggesting sites could flag or fact-check expertise. Yet expertise requires consensus. The viral YouTube film claiming COVID-19 death certificates were manipulated was by an anti-vaxx doctor, also a member of the Montana Health Board (26).

This study is not without limitations and invites extensions. We relied upon self-reports of media sources rather than objective logs. The data are cross-sectional, making it difficult to disentangle causality of whether exposure to poor vaccine and health information shapes hesitancy or a tendency to believe in conspiracies shapes information seeking. Although our study is representative complemented by focus groups, the sample size remains small in one country. Larger cross-national and longitudinal samples with multi-mode data gathering would be desirable.

## Materials and Methods

We commissioned Ipsos MORI to conduct a nationally representative online survey of 1,476 adults in the UK December 12 to 18, 2020 and five focus groups conducted between November 30 to December 7, 2020. Data deposition: Information about data, methods, and code are available at: medRxiv link

## Data Availability

Upon publication, processes to access data and code will be made available.

## Acknowledgments

Research Funding support has been provided by the ESRC’s UK in a Changing Europe programme, the Leverhulme Centre (Leverhulme Centre for Demographic Science) and ERC-2018-ADG-835079.

## Footnotes

*To whom correspondence may be addressed: Will Jennings and Melinda C. Mills

## Author contributions

WJ and MCM designed research and wrote the paper. GS, HW, VV, JG, DD, LW designed research and analysed data. All authors provided comments and approved the final draft.

The authors declare no competing interests.

This article contains supporting information online available at: medRxiv link

## References

1. M. C. Mills, D. Salisbury, The challenges of distributing COVID-19 vaccinations. EClinicalMedicine, 100674 (2020).

2. A. de Figueiredo, C. Simas, E. Karafillakis, P. Paterson, H. J. Larson, Mapping global trends in vaccine confidence and investigating barriers to vaccine uptake: a large-scale retrospective temporal modelling study. Lancet 396, 898–908 (2020).

3. M. C. Mills, et al., COVID-19 vaccine deployment: Behaviour, ethics, misinformation and policy strategies. R. Soc. (2020).

4. D. Devine, J. Gaskell, W. Jennings, G. Stoker, Trust and the Coronavirus Pandemic: What are the Consequences of and for Trust? An Early Review of the Literature. Polit. Stud. Rev., 147892992094868 (2020).

5. S. E. Kreps, D. L. Kriner, Model uncertainty, political contestation, and public trust in science: Evidence from the COVID-19 pandemic. Sci. Adv. 6, eabd4563 (2020).

6. P. M. Massey, Where Do U.S. Adults Who Do Not Use the Internet Get Health Information? Examining Digital Health Information Disparities From 2008 to 2013. J. Health Commun. 21, 118–124 (2016).

7. O. Oladejo, et al., Comparative analysis of the Parent Attitudes about Childhood Vaccines (PACV) short scale and the five categories of vaccine acceptance identified by Gust et al. Vaccine 34, 4964–4968 (2016).

8. J. D. Featherstone, R. A. Bell, J. B. Ruiz, Relationship of people’s sources of health information and political ideology with acceptance of conspiratorial beliefs about vaccines. Vaccine 37, 2993–2997 (2019).

9. F. J. Elgar, A. Stefaniak, M. J. A. Wohl, The trouble with trust: Time-series analysis of social capital, income inequality, and COVID-19 deaths in 84 countries. Soc. Sci. Med. 263, 113365 (2020).

10. D. Freeman, et al., Coronavirus conspiracy beliefs, mistrust, and compliance with government guidelines in England. Psychol. Med., 1–13 (2020).

11. A. Ehsan, H. S. Klaas, A. Bastianen, D. Spini, Social capital and health: A systematic review of systematic reviews. SSM - Popul. Heal. 8, 100425 (2019).

12. J. B. Dowd, et al., Demographic science aids in understanding the spread and fatality rates of COVID-19. Proc. Natl. Acad. Sci. 117, 9696–9698 (2020).

13. J. B. Dowd, P. Block, M. Jones, M. C. Mills, The Human Cost of Natural Herd Immunity. LCDS Blog (2020).

14. H. Seale, et al., Why do I need it? I am not at risk! Public perceptions towards the pandemic (H1N1) 2009 vaccine. BMC Infect. Dis. 10, 99 (2010).

15. A. Bish, L. Yardley, A. Nicoll, S. Michie, Factors associated with uptake of vaccination against pandemic influenza: a systematic review. Vaccine 29, 6472–84 (2011).

16. N. F. Johnson, et al., The online competition between pro- and anti-vaccination views. Nature 582, 230–233 (2020).

17. E. Hussein, P. Juneja, T. Mitra, Measuring Misinformation in Video Search Platforms: An Audit Study on YouTube. Proc. ACM Human-Computer Interact. 4, 1–27 (2020).

18. W. Jacobs, A. O. Amuta, K. C. Jeon, C. Alvares, Health information seeking in the digital age: An analysis of health information seeking behavior among US adults. Cogent Soc. Sci. 3, 1302785 (2017).

19. N. Smith, T. Graham, Mapping the anti-vaccination movement on Facebook. Information, Commun. Soc. 22, 1310–1327 (2019).

20. R. M. Anderson, C. Vegvari, J. Truscott, B. S. Collyer, Challenges in creating herd immunity to SARS-CoV-2 infection by mass vaccination. Lancet 396, 1614–1616 (2020).

21. C. H. Basch, P. Zybert, R. Reeves, C. E. Basch, What do popular YouTube videos say about vaccines? Child. Care. Health Dev. 43, 499–503 (2017).

22. G. Donzelli, et al., Misinformation on vaccination: A quantitative analysis of YouTube videos. Hum. Vaccin. Immunother. 14, 1654–1659 (2018).

23. D. Kahneman, Thinking Fast and Slow (Farrar, Straus and Giroux, 2011).

24. A. Barker, H. Murphy, Advertisers strike deal with Facebook and YouTube on harmful content. Financ. Times (2020).

25. A. Ohlheiser, Facebook and YouTube are rushing to delete “Plandemic”, a conspiracy-laden video. MIT Technol. Rev. (2020).

26. E. J. Dickson, Anti-vax doctor promotes conspiracy theory that death certificates falsely cite COVID-19. Roll. Stone (2020).

