## Supplementary Information for "Lack of trust and social media echo chambers predict COVID-19 vaccine hesitancy"

### Materials and Methods

#### Survey Design

We commissioned Ipsos MORI to conduct a nationally representative online survey of 1,476 adults in the UK December 12–18 2020. The survey asked respondents a series of questions about their perceptions of the COVID-19 pandemic including: various dimensions of trust in government, experts and the media, belief in conspiracy theories in general and specifically related to COVID-19, distrust of vaccines, knowledge about the coronavirus, scepticism towards the coronavirus and government restrictions ('lockdown scepticism'), the sources that people obtained their news information from, evaluations of the performance of government in handling the crisis and whether people were willing to be vaccinated for COVID-19.

Our survey was designed to investigate what might impact vaccine take-up or hesitancy. Broadly our measures were connected to trust across a range of arenas: from trust in government in general (including measures of mistrust and distrust), to trust in experts and information from the media, in addition to distrust in vaccines and general conspiracy beliefs. They also included perceptions of the threat posed by COVID-19 (to people personally, to their jobs/businesses, and to the country), and how well government was considered to be handling particular aspects of the crisis. We also examined how respondents consumed or shared information, and their use of 'vertical' (TV, radio, newspaper) or 'horizontal' (online, talking to people) sources for following news about politics or current affairs, as well as their use of specific social media platforms. We collected information on key demographic variables (age, gender, education, social grade, urban-rural, children in household), current voting intentions, and whether people had tested positive for, or believed they had been infected with, COVID-19.

#### Focus Group Design

We also ran five focus groups exploring themes of trust and COVID-19 November 30 to December 7 2020 with 29 participants across five locations in Bristol and Oldham, UK. A description of the focus group sample is shown in Table S1. One of the topics was whether people were willing to be vaccinated. We also asked to what extent they trusted the current government to manage the coronavirus crisis, how much they trusted information from the government, their views on conspiracy theories and stories circulating about COVID-19, the effectiveness of local lockdowns and the tier system, the balance between minimizing infections and keeping the economy going, and whether a vaccine is the only way the country can get 'back to normal'.

Table S1. Description of five focus groups, information collected between 30 November and 7 December 2020.

| Group | Location | Date | No participants | Voted in 2019 General Election | EU Referendum Vote | Age | Social Class | Ethnicity | Gender | Vaccine uptake profile |
|---|---|---|---|---|---|---|---|---|---|---|
| 1 | Bristol | 30.11.20 | 6 | Mix of political parties voted for (maximum x2 non-voters per group) | Remain | 35-54 | ABC1 | No quota – record for information purposes | Even mix of men and women | 1 yes<br>3 unsure<br>2 no |
| 2 | Oldham | 01.12.20 | 6 | | Leave | 35-54 | C2DE | | Even mix of men and women | 1 yes<br>1 unsure<br>4 no |
| 3 | Oldham | 02.12.20 | 5 | | Leave | 18-34 | ABC1 | | Even mix of men and women | 4 yes<br>1 not straight away |
| 4 | Oldham | 02.12.20 | 6 | | Remain | 18-34 | C2DE | | Even mix of men and women | 1 yes<br>5 no |
| 5 | Bristol | 07.12.20 | 6 | | Remain | 35-54 | ABC1 | | Even mix of men and women | 6 yes |

Notes: Derived from the National Readership Survey, ABC1 = [AB] high managerial, administrative or professional, [B] Intermediate managerial, administrative or professional, and [C1] supervisory, clerical and junior managerial, administrative or professional; C2DE = [C2] skilled manual workers, [D] semi and unskilled manual workers, and [E] state pensioners, casual or lowest grade workers, unemployed with state benefits only).

### Measurement and Methods

#### Survey measurement

Here we provide a full description of the survey variables used in this analysis, variables and coding and creation of composite indexes, referencing the original developers of those measures where relevant.

##### COVID VACCINE

Q1. To what extent, if at all, do you agree, or disagree, with the following statements?

1. A mass vaccination programme is the only way the country can return to normal
2. I trust the government to approve a safe and effective vaccine
3. I trust the government to develop an immunisation programme that will benefit everyone in our society
4. If a vaccine for COVID-19 were available to me, I would get it
5. If a vaccine for COVID-19 were available, I would get it for my child/children [if KIDS]
6. I would be willing to be tested for COVID-19 as part of a mass-testing programme
7. I would self-isolate for 14 days if advised to by the NHS app
8. I would self-isolate if I discovered that somebody I had come into contact with had tested positive for COVID-19
    a. Strongly agree
    b. Tend to agree
    c. Neither agree nor disagree
    d. Tend to disagree
    e. Strongly disagree
    f. Don’t know

##### SOCIAL TRUST

Q2. Generally speaking, would you say that most people can be trusted or that you need to be very careful in dealing with people?

1. Most people can be trusted
2. You need to be very careful
3. Don’t know

Source: [1]

##### TRUST IN HEALTH INSTITUTIONS

Q3. Here is a list of organizations. How much confidence do you have in each of the following: is it a great deal of confidence, quite a lot of confidence, not very much confidence or none at all?

1. The World Health Organisation
2. The NHS

    a. A great deal
    b. Quite a lot
    c. Not very much
    d. None at all
    e. Don’t know

Source: [1]

#### GOVERNMENT HANDLING OF COVID-19

Q4. To what extent, if at all, do you trust national government to do each of the following?

1. To handle the spread of COVID-19
2. To give reliable information about their handling of COVID-19
3. To treat people fairly and equally in their handling of COVID-19

   a. A great deal
   b. A fair amount
   c. Not very much
   d. Not at all
   e. Don't know

#### TRUST, MISTRUST, DISTRUST OF GOVERNMENT

Q4. To what extent do you agree, or disagree, with the following statements?

1. The government is honest and truthful [TRUST]
2. In general, the government usually does the right thing [TRUST]
3. The government acts unfairly towards people like me [DISTRUST]
4. The government usually ignores my community [DISTRUST]
5. I am usually cautious about trusting the government [MISTRUST]
6. I am unsure whether to believe the government [MISTRUST]
   a. Strongly agree
   b. Tend to agree
   c. Neither agree nor disagree
   d. Tend to disagree
   e. Strongly disagree
   f. Don't know

Source: [2]

#### CONSPIRIATORIAL BELIEFS

Q5. What do you think the likelihood is of each of the following being true or not true?
Please use a scale of 0 to 10, where 0 is certainly not true, and 10 is certainly true.

1. Many very important things happen in the world, which the public is never informed about
2. Politicians usually do not tell us the true motives for their decisions
3. Government agencies closely monitor all citizens
4. Events which superficially seem to lack a connection are often the result of secret activities
5. There are secret organisations that greatly influence political decisions
6. Much of what happens in the world today is decided by a small and secretive group of individuals
7. The real truth about the link between COVID-19 and 5G is being kept from the public
8. the government has a secret program that uses airplanes to put harmful chemicals into the air (often called "chemtrails")
9. The government is trying to cover up the link between vaccines and autism.
10. The coronavirus is a hoax

0 - Certainly not true
1
2

3
4
5 - About 50/50
6
7
8
9
10 - Certainly true
11. Don't know

Source: [3]

#### VACCINE CONSPIRACIES, DISTRUST & MISINFORMATION

Q6. Please rate the extent to which you agree or disagree with each of the following statements.

1. The coronavirus was bioengineered in a government-controlled lab in Wuhan, China
2. The spread of the coronavirus is a deliberate attempt to reduce the size of the global population
3. Powerful governments and business people deliberately devised COVID-19 in order to profit from a future vaccine
4. Tiny devices are placed in vaccines to track people
5. Vaccine safety data is often made up
6. People are being lied to about the effectiveness of vaccines
7. Data about the effectiveness of vaccines is often made-up
8. Vaccines are not harmful~~.~~
9. Hot temperatures kill the COVID-19 virus
10. The flu shot provides immunity to COVID-19
11. Hydroxychloroquine is <u>not</u> an effective treatment for COVID-19
12. The coronavirus is no worse than the seasonal flu

    a. Strongly disagree
    b. Disagree
    c. Somewhat disagree
    d. Neither agree nor disagree
    e. Somewhat agree
    f. Agree
    g. Strongly agree
    h. Don't know

Source: [4, 5, 6]

#### INFORMATION SOURCES

Q7. During the last month, on average how much time (if any) have you spent following news about politics or current affairs from each of these sources?

1. Television
2. Newspaper (including online)
3. Radio
4. Internet (not including online newspapers)
5. Talking to other people

    1. None, no time at all

2. At least once in the past month
3. Once a week
4. Several times a week
5. Once a day
6. Several times a day
7. Don’t know

Source: [7]

#### SOCIAL MEDIA

Q8. Which social media platforms, if any, have you visited in the past month?
Please tick all that apply.

1. Facebook
2. Twitter
3. Instagram
4. Reddit
5. YouTube
6. SnapChat
7. TikTok
8. Other (Please specify)
9. None of these
10. Don’t know

Q9. When using social media, how often, if at all, do you come across information about political issues?

1. Always
2. Most of the time
3. About half of the time
4. Sometimes
5. Never
6. Don’t know

Q10. During the last month, how often, if at all, have you done each of the following things…

1. Searched online for information about politics or current affairs
2. ‘Fact-checked’ an article about politics or current affairs (that is, checked its accuracy using other sources)
3. Posted information about politics or current affairs on social media (e.g. Facebook, Twitter)
4. Discussed politics or current affairs with people you know
5. Discussed politics or current affairs with people you don’t know
6. Seen other people discussing politics or current affairs online

    1. None, no time at all
    2. At least once in the past month
    3. Once a week
    4. Several times a week
    5. Once a day
    6. Several times a day
    7. Don’t know

#### TRUST, MISTRUST, DISTRUST IN MEDIA AND EXPERTS

Q11. Thinking about the traditional media (i.e. newspapers and television news, including their websites), how much if at all do you agree or disagree with the following statements?

1. I tend to trust information in the traditional media.
2. I tend to treat information from the traditional media cautiously, and check it against other sources, as much as possible.
3. I tend to believe that traditional media have vested interests which they don't reveal and are often unreliable in the information they provide.

    a. Strongly agree
    b. Tend to agree
    c. Neither agree nor disagree
    d. Tend to disagree
    e. Strongly disagree
    f. Don't know

Q12. To what extent, if at all, do you agree, or disagree, with the following statements?

1. Experts seem to disagree with each other as much as they agree
2. Experts have a good track record in predicting the future
3. Experts are poor at taking into account the views of people outside their field
4. Experts are key to solving the problems that our society faces
5. Evidence presented by experts needs to be treated with caution

    a. Strongly agree
    b. Tend to agree
    c. Neither agree nor disagree
    d. Tend to disagree
    e. Strongly disagree
    f. Don't know

#### LOCKDOWN SCEPTICISM

Q13. To what extent, if at all, do you agree, or disagree, with the following statements?

1. The government is placing too much emphasis on minimising infections from the coronavirus and not enough on keeping the economy going.
2. The government will eventually have to ease restrictions on our daily lives, even if that leads to more people catching the coronavirus.
3. The impact of the coronavirus is being exaggerated because most of the people dying would have died within a year or two anyway.
4. Young people and other groups at low risk of serious illness from the coronavirus should be free to go about their normal daily lives.
5. Eventually, the damage to people's lives from the lockdown will be greater than the health problems and fatalities caused by the coronavirus.
6. If you can prove you have recovered from the coronavirus, you should not be subject to restrictions on what you can do.
7. Local lockdowns are not effective in reducing spread of the coronavirus.
8. Local lockdowns wreck the economies of the areas affected by them.
9. I follow the rules, but a lot of other people aren't doing so.
10. Areas are being put into local lockdowns when they don't need to be.

11. Many deaths that are officially recorded as COVID-19 deaths are primarily due to other causes
    a. Strongly agree
    b. Tend to agree
    c. Neither agree nor disagree
    d. Tend to disagree
    e. Strongly disagree
    f. Don't know

Source: [8]

#### COVID-19 EXPERIENCE

Q14. Have you or any member of your household been furloughed or received financial help from the government in response to the coronavirus outbreak?

1. Yes
2. No
3. Don't know

Q15. Have you tested positive for, or believe that you have had, COVID-19?

1. Yes
2. No
3. Don't know
4. Prefer not to say

#### THREAT PERCEPTION

Q16. What level of threat, if any, do you think the coronavirus or COVID-19 poses to each of the following?

1. You personally
2. Your country
3. Your local area
4. Your job or business

    a. Very high threat
    b. High threat
    c. Moderate threat
    d. Low threat
    e. Very low threat
    f. Don't know

#### DEMOGRAPHIC AND POLITICAL CONTROLS

1. Vote intention
2. Leave/Remain support in the 2016 EU referendum
3. Age
4. Gender
5. Education
6. Region
7. Social class
8. Left-right self-placement

#### REFERENCES

[1] World Values Survey: http://www.worldvaluessurvey.org/

[2] Devine D, Gaskell J, Jennings W, Stoker G. 2020. 'Trust and the Coronavirus Pandemic: What are the Consequences of and for Trust? An Early Review of the Literature.' *Political Studies Review.* August 2020. doi:10.1177/1478929920948684

[3] Bruder M, Haffke P, Neave N, Nouripanah N, Imhoff R. 2013. 'Measuring individual differences in generic beliefs in conspiracy theories across cultures: conspiracy mentality questionnaire.' *Frontiers in Psychology* 4(225). doi:10.3389/fpsyg.2013.00225

[4] Jolley, DD, Douglas, KM. 2014. The effects of anti-vaccine conspiracy theories on vaccination intentions. *PloS One* 9(389177). doi:10.1371/journal.pone.0089177.

[5] Shapiro GK, Holding A, Perez S, Amsel R, Rosberger Z. 2016. 'Validation of the vaccine conspiracy beliefs scale.' *Papillomavirus Research* 2: 167-172. doi:10.1016/j.pvr.2016.09.001.

[6] Pickles K, Cvejic E, Nickel B, Copp T, Bonner C, Leask J, Ayre J, Batcup C, Cornell S, Dakin T, Dodd RH, Isautier JMJ, McCaffery KJ. 2020. 'COVID-19: Beliefs in misinformation in the Australian community.' doi: 10.1101/2020.08.04.20168583.

[7] British Election Study: https://www.britishelectionstudy.com/data-objects/panel-study-data/

[8] Sturgis P, Jackson K, Kuha J. 2020. 'Lockdown scepticism is part of the Brexit divide.' *LSE blog, June 8th, 2020.* https://blogs.lse.ac.uk/brexit/2020/06/08/20111/

### Methods

Table S3 provides a detailed description of the coding of all variables used in the analysis. The dependent variable of vaccine willingness is whether an individual agreed or disagreed with the statement “If a vaccine for COVID-19 were available to me, I would get it”, dichotomising the categories by coding respondents who said they strongly agreed or tend to agree (1) or strongly disagree, tend to disagree, neither or don’t know (0). The distribution of responses is plotted in Figure S1.

In our final analysis, we estimated logit regression models and present the odds ratios of willingness to be vaccinated. We also tried different specifications in the form of an ordered logit model and a partial proportional odds model that relaxed the proportional odds / parallel lines assumption and allowed the variables to vary with the point at which the categories of the dependent variable were dichotomized. Results were not markedly different and thus for ease of presentation opted for the logit model.

As noted in the main text, of our respondents, 49% strongly agreed they would get the vaccine, 22% indicated they tended to agree they would get it, 11% neither agreed or disagreed, 7% tended to disagree and 7% strongly disagreed. The combined share willing to get the vaccine (71%) is in line with results from other surveys, while the remaining 29% who are vaccine hesitant or worse (who either don’t agree or are unsure) presents a significant target group for policy-makers.

Figure S1. Summary statistics of the dependent variable of vaccine willingness (‘If a vaccine for COVID-19 were available to me, I would get it’). N = 1, 476

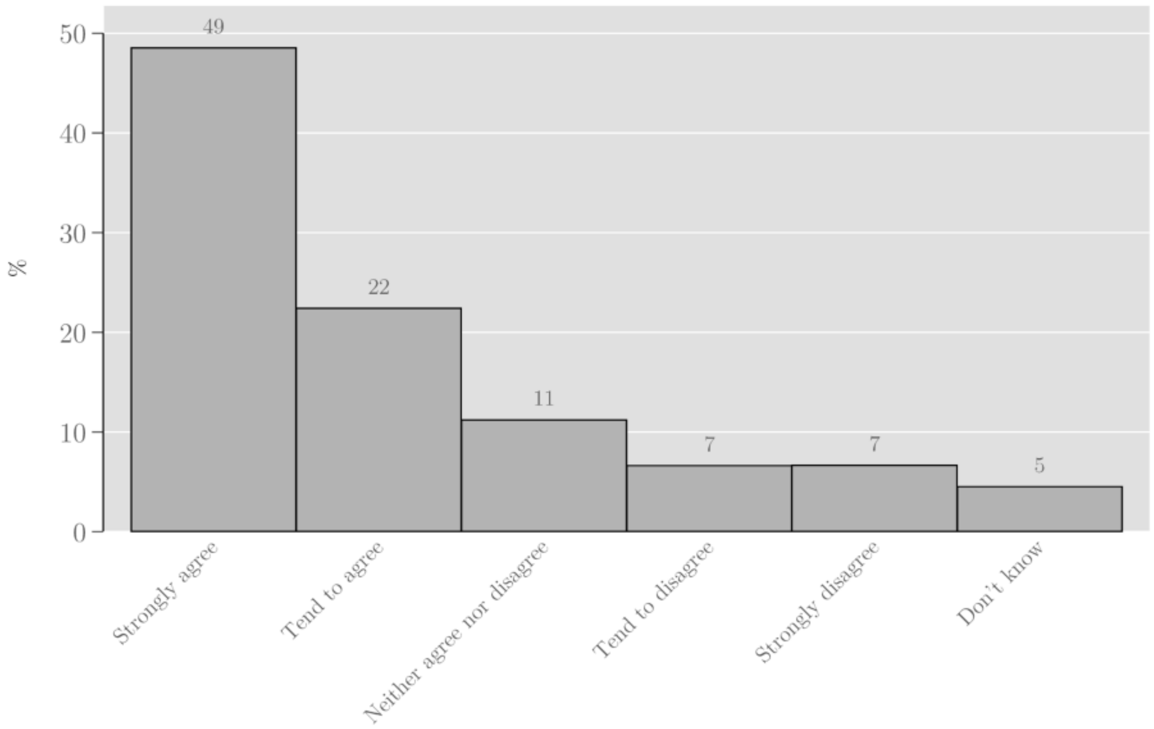

Most variables are rescaled to values between 0 and 1 (to facilitate direct comparison of effects) and where we used combined indices we calculate the additive index before rescaling. Dichotomous variables are used for yes/no questions (such as on whether respondents have visited particular social media platforms in the past month or have posted political content online). A number of demographic variables are also binary: whether respondents have tested positive for COVID-19 or believe they have suffered from the virus, gender (female) and education (degree or higher). We use a continuous variable for age, again rescaled from 0 to 1.

Table S2. Detailed coding of variables in the analysis

| Measure(s) | Coding | Question(s) |
|---|---|---|
| Social trust | 0 = You need to be very careful / Don't know<br>1 = Most people can be trusted | Q1 |
| Trust: government | 1 = Strongly disagree<br>2 = Tend to disagree<br>3 = Neither agree nor disagree<br>4 = Tend to agree<br>5 = Strongly agree<br><br>Additive index, rescaled: 0 to 1. | Q4 (1-2) |
| Mistrust: government | 1 = Strongly disagree<br>2 = Tend to disagree<br>3 = Neither agree nor disagree<br>4 = Tend to agree<br>5 = Strongly agree<br><br>Additive index, rescaled: 0 to 1. | Q4 (5-6) |
| Distrust: government | 1 = Strongly disagree<br>2 = Tend to disagree<br>3 = Neither agree nor disagree<br>4 = Tend to agree<br>5 = Strongly agree<br><br>Additive index, rescaled: 0 to 1. | Q4 (3-4) |
| Trust: health organisations (NHS, WHO) | 1 = None at all<br>2 = Not very much<br>3 = Quite a lot<br>4 = A great deal<br><br>Additive index, rescaled: 0 to 1. | Q3 (1-2) |
| Trust: media | 1 = Strongly disagree<br>2 = Tend to disagree<br>3 = Neither agree nor disagree<br>4 = Tend to agree<br>5 = Strongly agree<br><br>Rescaled: 0 to 1. | Q11 (1) |
| Trust: experts | 1 = Strongly disagree<br>2 = Tend to disagree<br>3 = Neither agree nor disagree<br>4 = Tend to agree<br>5 = Strongly agree<br><br>Additive index, rescaled: 0 to 1. | Q12 (2, 4) |
| Government handling of COVID-19 | 1 = None at all<br>2 = Not very much | Q4 (1-3) |

| | | |
|---|---|---|
| | 3 = Quite a lot<br>4 = A great deal<br><br>Additive index, rescaled: 0 to 1. | |
| Perceived personal threat of COVID-19 | 1 = Very low threat<br>2 = Low threat<br>3 = Moderate threat<br>4 = High threat<br>5 = Very high threat<br><br>Rescaled: 0 to 1. | Q16 (1) |
| Lockdown scepticism | 1 = Strongly disagree<br>2 = Tend to disagree<br>3 = Neither agree nor disagree<br>4 = Tend to agree<br>5 = Strongly agree<br><br>Additive index, rescaled: 0 to 1. | Q13 (1-11) |
| Conspiracy beliefs | 0 = Certainly not true<br>1<br>2<br>3<br>4<br>5 = About 50/50<br>6<br>7<br>8<br>9<br>10 = Certainly true<br><br>Additive index, rescaled: 0 to 1. | Q5 (1-5, 7-11) |
| Vaccine distrust | 1 = Strongly disagree<br>2 = Disagree<br>3 = Tend to disagree<br>4 = Neither agree nor disagree<br>5 = Tend to agree<br>6 = Agree<br>7 = Strongly agree<br><br>Additive index, rescaled: 0 to 1. | Q6 (5-7)<br><br>Q6 (8) scale inverted |
| COVID-19 misinformed | 1 = Strongly disagree<br>2 = Disagree<br>3 = Tend to disagree<br>4 = Neither agree nor disagree<br>5 = Tend to agree<br>6 = Agree<br>7 = Strongly agree<br><br>Additive index, rescaled: 0 to 1. | Q6 (9-12)<br><br>Q6 (11) scale inverted |
| Had COVID-19 | 0 = No, Don't know<br>1 = Yes | Q15 |
| Female | 0 = No<br>1 = Yes | |
| Age | Continuous variable, rescaled: 0 to 1. | |
| Graduate | 0 = NVQ3 or lower<br>1 = Degree or higher qualification | |

| | | |
|---|---|---|
| Supports Conservative party | 0 = No (voting intention for another party or no voting intention)<br>1 = Voting intention for Conservative Party | |
| Information sources: online | 1 = None, no time at all<br>2 = At least once in the past month<br>3 = Once a week<br>4 = Several times a week<br>5 = Once a day<br>6 = Several times a day<br><br>Rescaled: 0 to 1. | Q7 (4) |
| Information sources: people | 1 = None, no time at all<br>2 = At least once in the past month<br>3 = Once a week<br>4 = Several times a week<br>5 = Once a day<br>6 = Several times a day<br><br>Rescaled: 0 to 1. | Q7 (5) |
| Information sources: traditional | 1 = None, no time at all<br>2 = At least once in the past month<br>3 = Once a week<br>4 = Several times a week<br>5 = Once a day<br>6 = Several times a day<br><br>Rescaled: 0 to 1. | Q7 (1-3) |
| Social media: Facebook | 0 = No<br>1 = Yes (visited in the past month) | Q8 (1) |
| Social media: Twitter | 0 = No<br>1 = Yes (visited in the past month) | Q8 (2) |
| Social media: Instagram | 0 = No<br>1 = Yes (visited in the past month) | Q8 (3) |
| Social media: Reddit | 0 = No<br>1 = Yes (visited in the past month) | Q8 (4) |
| Social media: YouTube | 0 = No<br>1 = Yes (visited in the past month) | Q8 (5) |
| Social media: SnapChat | 0 = No<br>1 = Yes (visited in the past month) | Q8 (6) |
| Social media: TikTok | 0 = No<br>1 = Yes (visited in the past month) | Q8 (7) |
| Posted political content online | 0 = No<br>1 = Yes (posted information about politics or current affairs on social media in the past month) | Q10 (3) |
| Fact-checked an article online | 0 = No<br>1 = Yes (‘fact-checked’ an article about politics or current affairs in the past month) | Q10 (2) |

### Additional Results and Sensitivity Analyses

#### Survey Results and Additional Analyses

##### Bivariate logit regression

We first estimated bivariate logit regressions of willingness to get the vaccine on our predictors. This enabled us to understand the relationship between each of the measures and vaccine uptake. Figure S2 plots the odds of willingness to get the COVID-19 vaccine by variable (with a log scale used for the odds ratio on the x-axis). To interpret these effects, where the odds ratio exceeds 1.0 (marked by the red vertical line), this indicates the predictor is associated with a greater willingness to be vaccinated, and where it is lower than 1.0, this indicates it is associated with a lower willingness to receive the vaccine.

**Figure S2.** Bivariate logit regression of vaccine willingness, odds ratios

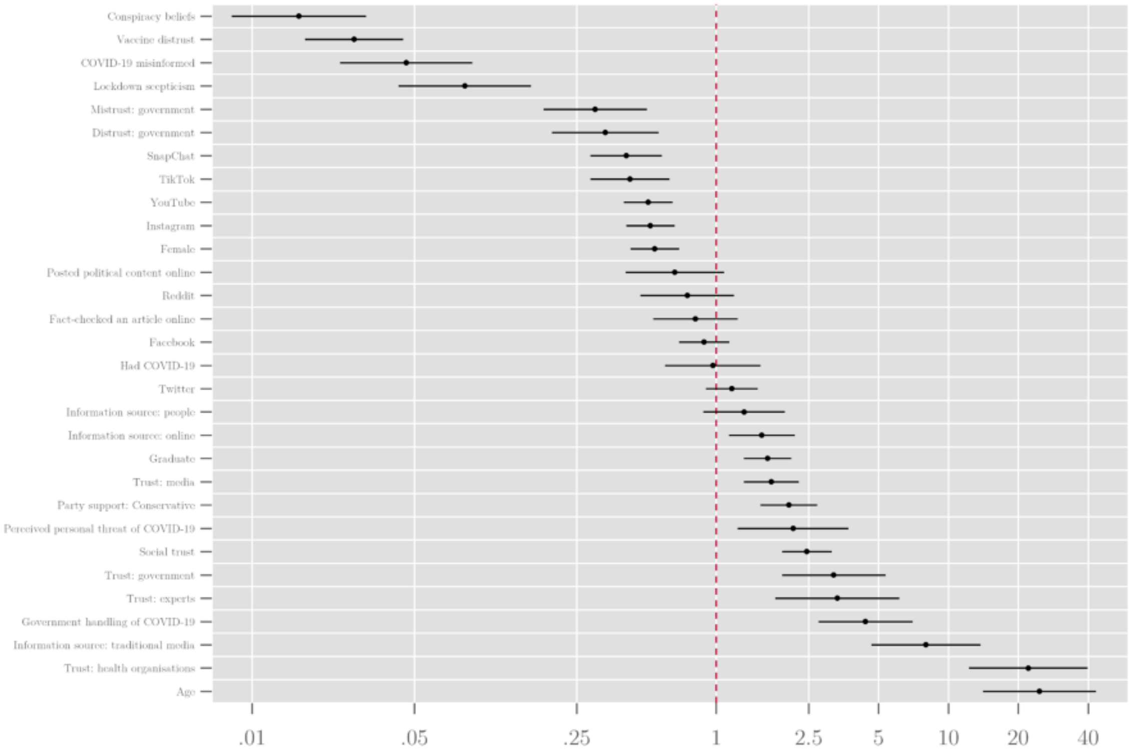

Of those factors that *increase* the likelihood of vaccine willingness, **age** and **trust in health organisations (i.e. the NHS and the WHO) had the strongest effect**. The odds ratio of just over 20 means that the oldest respondents are over twenty times more likely to expressing willingness to get the vaccine as the youngest. Similarly, someone with a high level of trust in health organisations is around twenty times more likely to be willing to be vaccinated than someone with the lowest level of trust. The next largest positive association is for people who consume a large amount of information from traditional media, followed by positive evaluations of government handling of the COVID-19 crisis, trust in experts and government, social trust, perceived personal threat from COVID-19, support for the governing Conservative Party, trust in information from the media, those with a degree or above, and those who consume a large amount of information online.

Of those factors that *decrease* the likelihood of willingness to get the vaccine, conspiracy beliefs have the largest effect, followed by **distrust of vaccines** (our battery of four questions designed to measure distrust: 'vaccine safety data is often made up', 'people are being lied to about the effectiveness of vaccines', 'data about the effectiveness of vaccines is often made-up', 'vaccines are not harmful'), belief in COVID-19 misinformation, and 'lockdown scepticism' (Sturgis et al. 2020). General mistrust and distrust in government are associated with odds of being willing to get the vaccine that are around three times lower. Users of Instagram, YouTube, Snapchat and TikTok are all less likely to express willingness to be vaccinated, as are women.

Because vaccine distrust is proximate to our outcome variable (willingness to be vaccinated against COVID-19), we also estimated the model excluding it as a predictor, as shown in Figure S3. This had minimal impact on the results, indicating that effects of other attitudinal and behavioural predictors are robust to its inclusion.

**Figure S3.** Multivariate logistic regression of vaccine willingness (vaccine distrust excluded), odds ratios

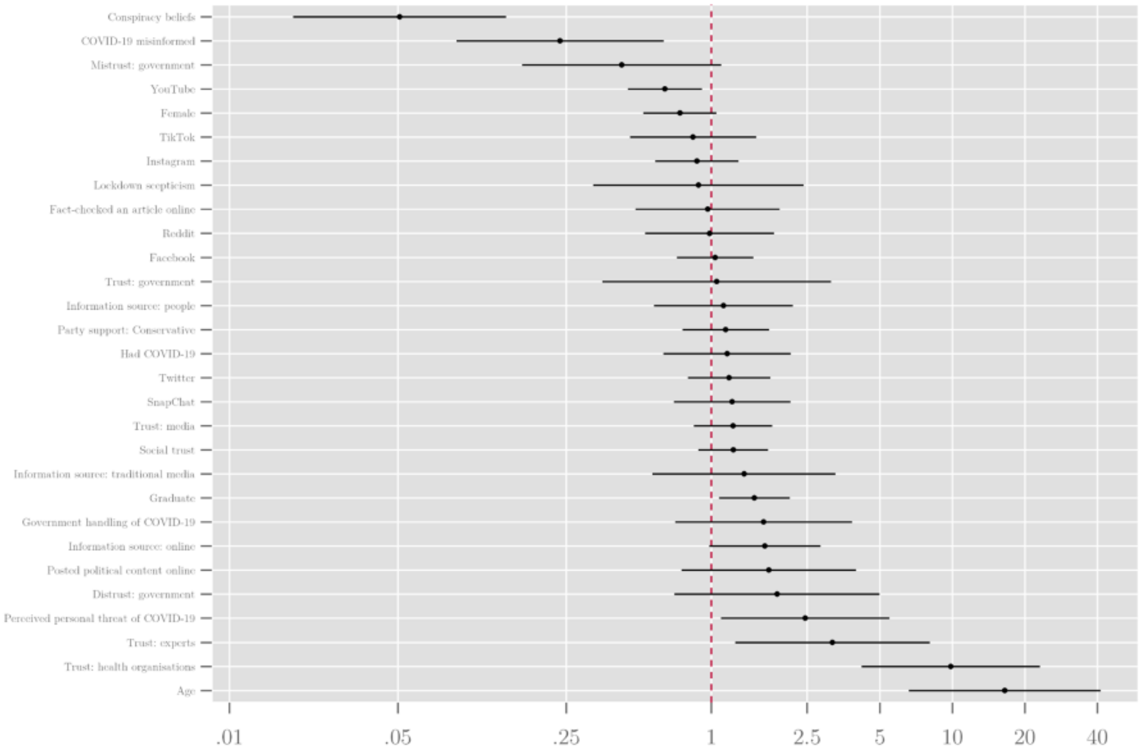

#### Multivariate logit regression, predictors controlling for demographics

We next proceeded to estimate logit regression models of willingness to get vaccinated, controlling for demographics and attitudinal factors. Figure S4 presents individual models with specific sets of predictors controlling for demographics. These broadly confirm the findings from the bivariate regression. It is noticeable that attitudinal predictors typically have larger effects than demographic predictors. The beliefs that individuals hold tend to be a stronger guide to whether or not they are vaccine hesitant than their demographic characteristics.

Figure S4. Multivariate logistic regression models of vaccine willingness, odds ratios

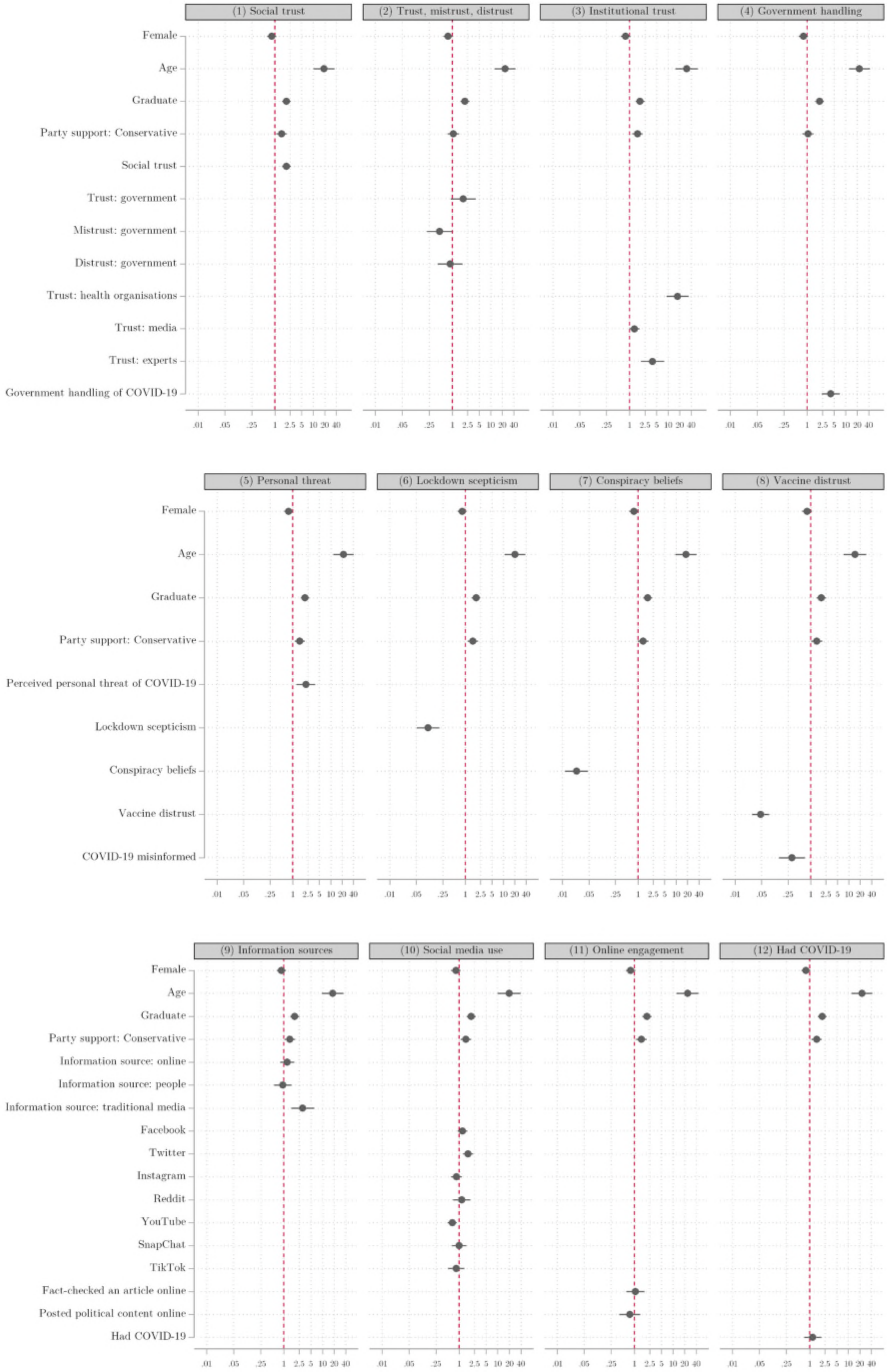

#### Focus Groups and Additional Analyses

Table S3 provides a description of the focus group participants by their vaccine uptake opinion and other key factors including location, social class, whether they were furloughed and their trust profile. Of the 29 participants in total:

- 14 stated that they would take the vaccine, but 1 not straight away;
- 11 would not take the vaccine;
- 4 were unsure.

Table S3. Description of focus group participants by vaccine uptake opinions

| Vaccine uptake | Location | Social grade | Furlough? | Trust profile pre-group |
|---|---|---|---|---|
| Yes | 7 Bristol; 6 Oldham | 1 A; 4 B; 6 C1; 2 C2 | 1 furloughed; positive views of furlough mentioned | 6 trusters; 3 mistrusters; 4 distrusters |
| Not straight away | 1 Oldham | B | No mention | 1 mistruster |
| Unsure | 3 Bristol; 1 Oldham | 2 C1; 1 C2; 1 B | 2 furloughed | 2 trusters and 2 mistrusters |
| No | 9 Oldham; 2 Bristol | 2 C1; 6 C2; 2 D; 1 E | 1 furloughed | 3 trusters; 4 mistrusters; 4 distrusters |

*Note: social grade categories, derived from the National Readership Survey (A = high managerial, administrative or professional; B = Intermediate managerial, administrative or professional; C1 = supervisory, clerical and junior managerial, administrative or professional; C2 = skilled manual workers; D = semi and unskilled manual workers; E = state pensioners, casual or lowest grade workers, unemployed with state benefits only).*

Those who said they **would** take the vaccine were more likely to have stated that they trusted the government's handling of the pandemic. Interestingly, there is acknowledgement in this group of the inconsistencies, and even referring to incompetence, but an implicit (and sometimes explicit) trust that the government are trying their best or to do the right thing. Indeed, those in this group were the ones most likely to mention positive attitudes about the furlough scheme, possibly associating this with benevolence. Similarly, in assessing the balance between protecting lives and supporting the economy, they recognised the difficulty that the government faced.

These participants were also more likely to see the government as having followed the science; though they were split on whether the virus was a natural occurrence or man-made, with some expressing doubt over the validity of COVID deaths. They seemed to implicitly trust the science and vaccine approval processes, recognising the extraordinary effort that has gone into getting to that point. They also understood the (mRNA – although no one explicitly mentioned the term) vaccine to be a relatively new kind of technology. There was also broad recognition of the need for a vast majority of people to get vaccinated. The main reasons stated for their decision to have the vaccine was to protect their families and or as their civic duty to protect society. They saw it as the only way back to some form of normality.

The participant who would **not have the vaccine straight away** expressed his own theory of the man-made origin of the coronavirus and then asserted the need to see proof of it working before accepting to take it.

Those who were **unsure** were mainly nervous about the rapidity of the vaccine development process, identifying the need for more testing. They did not feel that a vaccine was the only way back to normality, largely attributed to mixed interpretations around the notion of '**herd immunity**'. This group also expressed a lack of trust in the information from the government about the crisis, citing inconsistencies on how **COVID deaths were recorded** as justification. They also expressed scepticism or real uncertainty around theories over the origin of the virus, saying it is very difficult to know what to believe. Finally, one participant mentioned being hesitant around the vaccine because of the idea that some form of vaccine passport would be required in order to return to normality. Another outlined the different efficacies of the various vaccines, suggesting the government had purchased more of the less effective vaccine.

Those who stated that they **would not** get vaccinated were more likely to have stated that they trusted the government's response to the pandemic the least, citing as justification the perception that there is 'one rule for us, another for them'; scepticism around reported COVID death figures; and the unfairness (politicisation) and inconsistencies of the tier system. Participants who declared that they would not get the vaccine pointed to the policy confusion, scandals over PPE (personal protective equipment), schools, the Prime Minister not attending COBRA[1] meetings, perceived corruption and policy leaking to newspapers as evidence of generally untrustworthy behaviour. Furthermore, this group tended not to trust information they received from the government on the crisis. They were also less likely to believe the government had 'followed the science' (a term that was brought up in discussions) throughout.

A common thread amongst these participants was their view that the government put too much emphasis on lockdown measures at the expense of the economy. There participants mentioned the longer-term economic fallout on livelihoods, the politicised nature of the tier system, which they saw as punishment for Andy Burnham (Mayor of the Northern city of Manchester) standing up to the government and strong favouritism for London over the North. The majority of people (8) who would refuse the vaccine either believed the virus was man-made, or were willing to keep an open mind to this possibility. This was because they identified the uneven effects of the virus on different population groups as some sort of targeting which they perceived as unnatural and as a form of population control. None of them believed the vaccine was the only way back to normality. In fact, they offered either some adapted understanding of herd immunity, or that the virus was not as deadly as described (linked to scepticism of registered deaths) concluding that most people do not need a vaccine. Similarly, in justifying their decision not to get a vaccine, they highlighted that the vaccine process had been rushed, not enough testing undertaken, and the existence of unknown side-effects. One compared it to the thalidomide scandal of the late 1950s as an example of what goes wrong with untested medicine. The assessments did not take into account the fact that in order to be effective in a population, a vaccine needs to be administered to a sufficient percentage of the population. Instead, people believed that those who were most vulnerable to COVID-19 should potentially receive the vaccine, but as they did not find themselves in an at-risk category, they would not need a vaccine. Overall, this group concluded that the unknown possible side-effects from vaccine were a greater risk than the possible death or long-term effects of COVID.

[1] COBRA is a cross-departmental committee that comes together to respond to national emergencies in the UK and stands for Cabinet Office Briefing Rooms (COBR), often in meeting room A. Its aim is to make fast, effective decisions in a crisis and coordinate the response of the central government.
